# Alternative per-protocol estimates: secondary analyses of data from the Balanced randomised controlled trial

**DOI:** 10.1101/2024.10.01.24314731

**Authors:** Jim Young, Tim Short, Luzius A Steiner, Salome Dell-Kuster

**Affiliations:** Department of Epidemiology, Biostatistics and Occupational Health, Faculty of Medicine, McGill University, Montreal, Canada; Department of Anaesthesia and Perioperative Medicine, Auckland City Hospital, Auckland, New Zealand; Department of Anaesthesia, University of Auckland, Auckland, New Zealand; Clinic for Anaesthesiology, Intermediate Care, Prehospital Emergency Medicine and Pain Therapy, Department of Acute Medicine, University Hospital of Basel, Basel, Switzerland; Department of Clinical Research, University of Basel, Basel, Switzerland

**Keywords:** BIS, bispectral index, cognitive dysfunction, depth of anaesthesia, drug titration paradox, postoperative delirium, postoperative mortality

## Abstract

**Background:** The Balanced trial was designed to answer the question of whether anaesthetic depth affects postoperative mortality when a vulnerable patient undergoes major surgery. Patients were recruited between 2012 and 2017 at 73 centres in seven countries. In the intention-to-treat analysis (n=6644), there was no significant difference in one year mortality between patients randomised to surgery under deep (target BIS 35) or light anaesthesia (target BIS 50). However the separation between randomised groups was only 8.4 BIS units and the trial was criticised for being underpowered.

**Methods:** In a secondary analysis of this trial’s data, we made alternative per-protocol estimates designed to improve the power of the trial. We added an additional covariate – each patient’s deviation from target BIS – to the original analysis, statistically recreating the desired separation of 15 BIS units between randomised groups. We used multiple imputation to recover missing BIS values. We also assessed whether a proportional hazards Cox model was appropriate for the analysis of one year mortality.

**Results:** Our alternative per-protocol estimates did not differ materially from the original per-protocol estimate. The gain in precision through using all intent-to-treat patients for our per-protocol estimates was offset by the additional variance introduced when modelling missing BIS values. When modelling missing BIS, we found regional differences: in China, the separation between randomised groups was far higher (13.6 BIS units) than in any other region. Estimates and plots assessing proportional hazards suggested increasing late mortality under deep anaesthesia, most notably in China.

**Conclusion:** Our hypothesis is that deep anaesthesia in the Balance trial led to higher postoperative delirium, which in turn led to an increase in late mortality. In future trials, patients should be followed for more than a year and cause of death recorded.

**Key points:**

- We added an additional covariate – each patient’s deviation from their target BIS – to the original analyses of data from the Balanced trial, statistically recreating the desired separation of 15 BIS units between randomised groups.
- Our alternative per-protocol estimates for the effect of deep anaesthesia on one year mortality did not differ materially from the original per-protocol estimate.
- In China, the separation between randomised groups was far higher (13.6 BIS units) than in any other region (at most 7.7 BIS units).
- Estimates and plots assessing proportional hazards suggested increasing late mortality under deep anaesthesia, most notably in China.
- Our hypothesis is that deep anaesthesia in the Balance trial led to higher postoperative delirium, which in turn led to an increase in late mortality.

## Introduction

To an outsider it seems remarkable that controversy surrounds the risks of deep anaesthesia, given the fundamental nature of this question to anaesthesiology. Detrimental associations have been reported in observational data,^1^ leading to fierce debate: “many anesthesia clinicians now believe dogmatically that deep hypnosis during anesthesia is injurious, and this belief has become entrenched in anesthesiology and critical care lore and literature, despite the lack of robust corroborative evidence and even some compelling contradictory evidence.”^2^ The Balanced trial was designed to end the controversy.^3^ In a Herculean effort, 6644 older patients undergoing major surgery were randomised to general anaesthesia with bispectral index (BIS) values either between 30-40 (BIS 35 group) or between 45-55 (BIS 50 group).^4^ The investigators concluded that one year mortality did not differ between the two groups.

Unfortunately results from the trial did not end the debate. Scepticism was reflected in the titles of many editorials: “Deep anaesthesia and poor outcomes: the jury is still out”, “Deep anaesthesia and postoperative death: Is the matter resolved?”, “Achieving balance with power: lessons from the Balanced Anaesthesia Study”.^5-7^ Those commenting identified a lack of power as a key issue: “the sample size was inadequate to answer the trial’s primary question.”^7^ However, the Balanced trial was realistically about as large as it could have been, a point implicitly conceded by the conclusion that approaches other than a conventional randomised trial might be needed to answer the question.^6-8^

In a letter to The Lancet, we suggested an alternative per-protocol estimate that might improve the power of the trial.^9^ This report documents our efforts to make more precise estimates of the effect of deep anaesthesia on one year mortality using data from the Balanced trial and various forms of covariate adjustment. During the process of acquiring these data, a delirium sub-study of the Balanced trial was published, showing greater postoperative delirium – and worse one year cognitive function – in those randomised to deep anaesthesia.^10^ This only added to the controversy because of apparent discrepant findings between the parent trial and its sub-study, with deep anaesthesia having no significant effect in the former but a detrimental effect in the latter.^11^ We subsequently realised the results of our secondary analyses offered an explanation for this discrepancy and a hypothesis for how deep anaesthesia did indeed affect mortality in the Balanced trial; a hypothesis that deserves further study.

## Methods

### Ethics

A protocol for this study and a statistical analysis plan (Appendix A) were approved by the New Zealand Health and Disability Ethics Committees (EXP13559; H Walker, Chair; 15 November 2022) and by the Research Review Committee Te Toka Tumai Auckland (A+9688; M-A Woodnorth, Manager, Research Office; 16 December 2022). Data sharing was subject to a data management plan and a formal signed agreement (12 May 2023). The Balanced trial was registered with the Australian New Zealand Clinical Trials Registry (ACTRN12612000632897).

### Patient population

In the Balanced trial, high-risk older patients undergoing major surgery were enrolled between December 2012 and December 2017 in 73 centres across 7 countries.^4^ Patients had to be 60 years or older, with significant comorbidity (American Society of Anaethesiologists, ASA, physical status 3 or 4), having surgery with expected duration of more than two hours, and with an anticipated hospital stay of at least two days. Patients received volatile anaesthetic-based general anaesthesia either with or without major regional anaesthesia.

The Balanced trial was designed as an explanatory trial. The aim of the trial was “to definitively answer the question of causality and whether titrating anaesthetic depth makes a difference to patient outcome in a vulnerable patient group”.^3^ However the trial was analysed as a pragmatic trial, testing “whether an intervention to control depth within tight, predetermined parameters is effective”.^12^ This is not the same as answering the causal question of whether anaesthetic depth influences patient outcome. Per-protocol estimates are more useful for inferring causality than the intent-to-treat estimate, although best made using all patients in the trial to maximise precision and avoid bias.^13^

Non-compliance with the trial protocol reduced the difference in anaesthetic depth between the randomised groups.^4^ Most patients in the BIS 50 group had a BIS below target (median 47, interquartile range 44 to 51) while most patients in the BIS 35 group had a BIS above target (median 39, interquartile range 36 to 42). This meant that the intention-to-treat analysis was based on a between group BIS separation of only 8.4 units, well below the target of 15. In the original per-protocol analysis, patients were excluded if their achieved BIS was more than five units from target.^4^ This reduced the number of patients, from 6626 in the intent-to-treat analysis to 4060, a reduction of nearly 40%. As a result, the estimate of the effect of anaesthetic depth on one year mortality was of a lower precision in the per-protocol analysis than in the intent-to-treat analysis. Excluding these patients increased the separation between treatment groups, from 8.4 to 12.3 BIS units, but still less than the target of 15.

Our population of interest was the intent-to-treat population of the Balanced trial. We used this population to make a per-protocol estimate of the effect of anaesthetic depth by adding an additional covariate – each patient’s deviation from target BIS (35 or 50 depending on their randomised group) – to the original intent-to-treat analysis. Adding this covariate provides an adjusted estimate of the effect of anaesthetic depth for a patient with no deviation from target, under the assumption of an approximately log linear association between deviation from target BIS and outcome. In a statistical sense, this recreates the desired separation between randomised groups of 15 BIS units. It allows a per-protocol estimate to be made using all patients – with therefore no loss of precision – and, if a lower BIS is associated with increased mortality, adding this covariate might even increase the precision of the estimate.^14^

### Primary and secondary outcomes

The primary outcome for this study was one year mortality; the same primary outcome as in the Balanced trial. Secondary outcomes were a subset of those reported in the Balance trial: myocardial infarction, cardiac arrest, pulmonary embolism, stroke, sepsis, surgical site infection, and unplanned ICU admission. Our analyses of secondary outcomes are described in Appendix B.

### Statistical analyses

The original estimate of the effect of anaesthetic depth on one year mortality was made using a Cox model stratified by region with randomised group as the sole independent variable. Our base model was this Cox model with deviation from target BIS as an additional covariate. We then added other covariates – for other reasons.

First we added a covariate, age at surgery, to our base model to further improve precision. Analyses of randomised trials typically do not use prognostic covariates. However including covariates strongly associated with outcome will increase the precision of estimates of a treatment effect, even in non-linear models such as the Cox model.^15^

Second – to reduce bias – we then added two additional covariates (preoperative Charlson comorbidity index and preoperative WHODAS score) to the base model plus age. Mean achieved BIS can be thought of as a measure of adherence to the trial protocol. From this perspective, including the deviation from target BIS in a regression model for an outcome could erode the protection from confounding afforded by randomisation.^13^ Because mean achieved BIS is measured after randomisation, there is the possibility of confounding between deviation from target BIS and outcome, as in an observational study. Anaesthetists might be reluctant to approach the BIS target with patients they perceive as particularly frail. The solution is to include baseline covariates that could be associated with both deviation from target BIS and outcome.^13^

Finally we added a time dependent treatment variable to our base model to assess whether a proportional hazards Cox model was appropriate for these data. We used two simple ways to investigate non-proportional hazards: (1) adding a time dependent term to a Cox model to represent the interaction between randomised group and (typically log) time; and (2) transformed survival curves showing log minus log survival plotted against log time.^16^

Note that adding covariates to a non-linear model, like the Cox model, changes the treatment effect being estimated from a marginal effect to a conditional effect. The conditional effect is the average effect of shifting individuals with specific covariate values from light to deep anaesthesia.^17^ We centred all our covariates about mean values (age 72, Charlson comorbidity index 7.0, WHODAS score 20, log days to mortality 4.7). The conditional treatment effect then estimated is one appropriate for patients with these mean covariate values.

The only problem was mean achieved BIS value was missing for 3% of the patients in the intent-to-treat population.^4^ We replaced missing BIS values by multiple imputation. The assumption behind this step is that missing BIS values can be predicted by information recorded about patients prior to surgery. We assessed a variety of imputation models, without reference to outcome data, for their ability to minimise the variance between multiple imputed data sets. When imputing missing BIS, an imputation model ought to contain: (1) variables that are associated with whether BIS is missing; (2) variables that are associated with the value of BIS itself; and (3) any variables that will be subsequently included in an analytic model.^18^ Having selected a model, we then created a single sample of 200 multiply imputed data sets. This sample was used for all estimates, using Rubin’s rule for combining point estimates from multiple imputed data sets.^19^

Our analyses were carried out in SAS 9.4 TS Level 1M5 (procedures PHREG, MI and MIANALYZE). In contrast to the original analysis, we report hazard ratios (HR) with light anaesthesia as the reference – for two reasons. First, most meta-analyses summarising past trials and observational studies have taken this approach,^1,20-22^ although not all.^8,23^ Second, clinical concern is that deep anaesthesia will increase mortality. It is important to present results in a way that is consistent with other literature and with underlying clinical concern, if comparisons are then going to be made with that literature and results translated into implications for clinical practice. The hazard ratio scale is not symmetric, so that perceptions of point and interval estimates may change if estimates are inverted.

## Results

### Multiple imputation modelling

When modelling missing BIS, we found regional differences in mean achieved BIS (Table 1). In China, patients were closer to target BIS, in both randomised groups, than in any other region. The net effect was a separation between randomised groups in China of 13.6 BIS units, far higher than the next highest separation (7.7 BIS units in USA).

**Table 1.** Median mean achieved BIS in the intent to treat population by randomised group and region.

| Region | Light anaesthesia<br>(target BIS 50) | Deep anaesthesia<br>(target BIS 35) | Separation between<br>randomised groups |
| --- | --- | --- | --- |
| Australia | 46.2 | 39.0 | 7.2 |
| China | 51.3 | 37.7 | 13.6 |
| New Zealand | 46.5 | 39.0 | 7.5 |
| UK and Europe | 45.4 | 38.3 | 7.0 |
| USA | 48.2 | 40.5 | 7.7 |
| Overall | 47.2 | 38.8 | 8.4 |

Our final imputation model used full Markov Chain Monte Carlo imputation assuming multivariate normality for imputation variables, with each randomised group modelled separately. The imputation model for each randomised group included continuous variables age at surgery, preoperative Charlson comorbidity index, preoperative WHODAS 2.0 score and achieved mean BIS; and indicator variables for large and small hospitals (with mid-sized hospitals as the reference), for the regions China, UK and Europe, and USA (with Australia and New Zealand combined as the reference) and for ASA physical status 4 (with a lower status as the reference).

### Per-protocol estimates

The per-protocol estimate for the effect of deep anaesthesia on one year mortality, with adjustment for deviation from target BIS, was hazard ratio (HR) 1.16, 95% confidence interval (CI) 0.93 to 1.46; almost identical to the original per-protocol estimate (HR 1.16, 95% CI 0.92 to 1.46). The estimate with further adjustment to improve precision was HR 1.19, 95% CI 0.94 to 1.49. The estimate with further adjustment to reduce bias was HR 1.18, 95% CI 0.94 to 1.48.

There is no material difference between all four per-protocol estimates (Figure 1). All four are consistent with the 20% increase in mortality estimated from observational studies;^1^ the increase assumed in the sample size calculation for the Balanced trial.^3,4^

**Figure 1.**
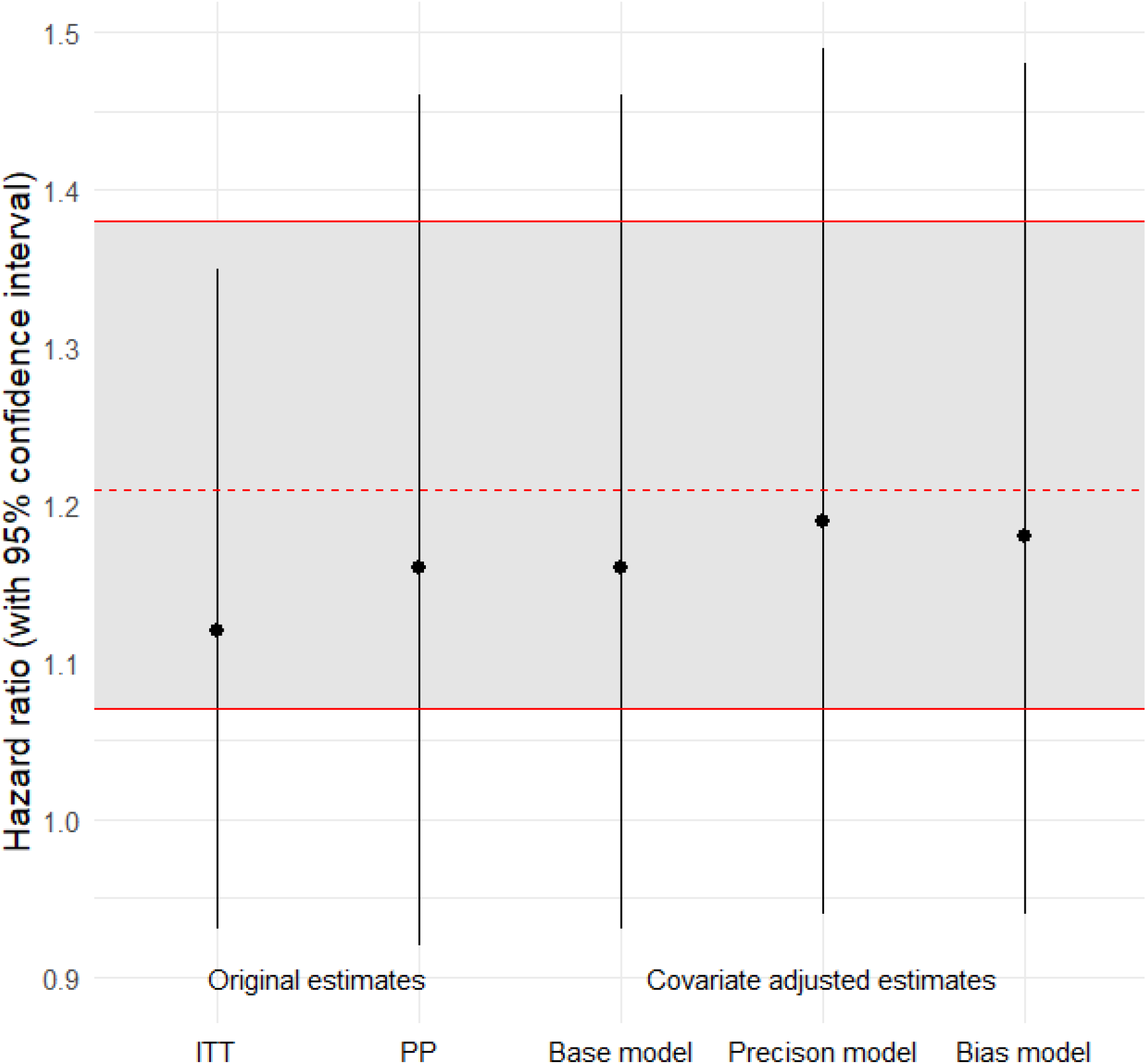
Cox model estimates of the effect of deep anaesthesia on one year mortality. The original estimates are from the original intent-to-treat (ITT) and per-protocol (PP) analyses.^4^ The covariate adjusted estimates are from this study: the base model is the original Cox model plus deviation from target BIS; the precision model is the base model plus age at surgery; the bias model is the base model plus age at surgery, preoperative Charlson comorbidity index and preoperative WHODAS score. The shaded band shows the point estimate and 95% confidence interval from a meta-analysis of observational studies;^1^ a 20% increase was used to calculate a sample size for the Balanced trial.^3,4^

### Proportional hazards

The per-protocol estimate for the effect of anaesthetic depth on one year mortality, with adjustment for deviation from target BIS and a time dependent interaction term, was HR 1.16, 95% CI 0.93 to 1.46. The estimate of the time dependent interaction suggests an increasing trend in the hazard over time (HR 1.07, 95% CI 0.93 to 1.23).

To illustrate, consider a model with two hazard ratios, one for events during the first 30 days and one for events after that. Such a model is consistent with the design of the trial, with comparisons between randomised groups planned at 30 days and one year.^3^ Results for this model, with adjustment for deviation from target BIS, suggested no risk of increased mortality in the first 30 days (HR 0.94, 95% CI 0.59 to 1.53) but potentially an increasing risk of mortality after that (HR 1.21, 95% CI 0.95 to 1.54).

A proportional hazard ratio will lead to parallel curves for the two randomised groups in plots of log minus log survival against log time.^16^ Such plots suggest the curves are not parallel (Figures 2 and 3). This is clear in the plot for China: the curve for deep anaesthesia crosses the curve for light anaesthesia (Figure 2). Curves for deep anaesthesia cross for Australia too or steepen over time for the USA (Figure 3). For the UK and Europe and for New Zealand, curves for deep anaesthesia might be steeper towards the end of the trial but it is not certain and longer follow-up of trial participants is needed. These curves suggest that one year might have been too early to see a clear difference in mortality between the two randomised groups.

**Figure 2.**
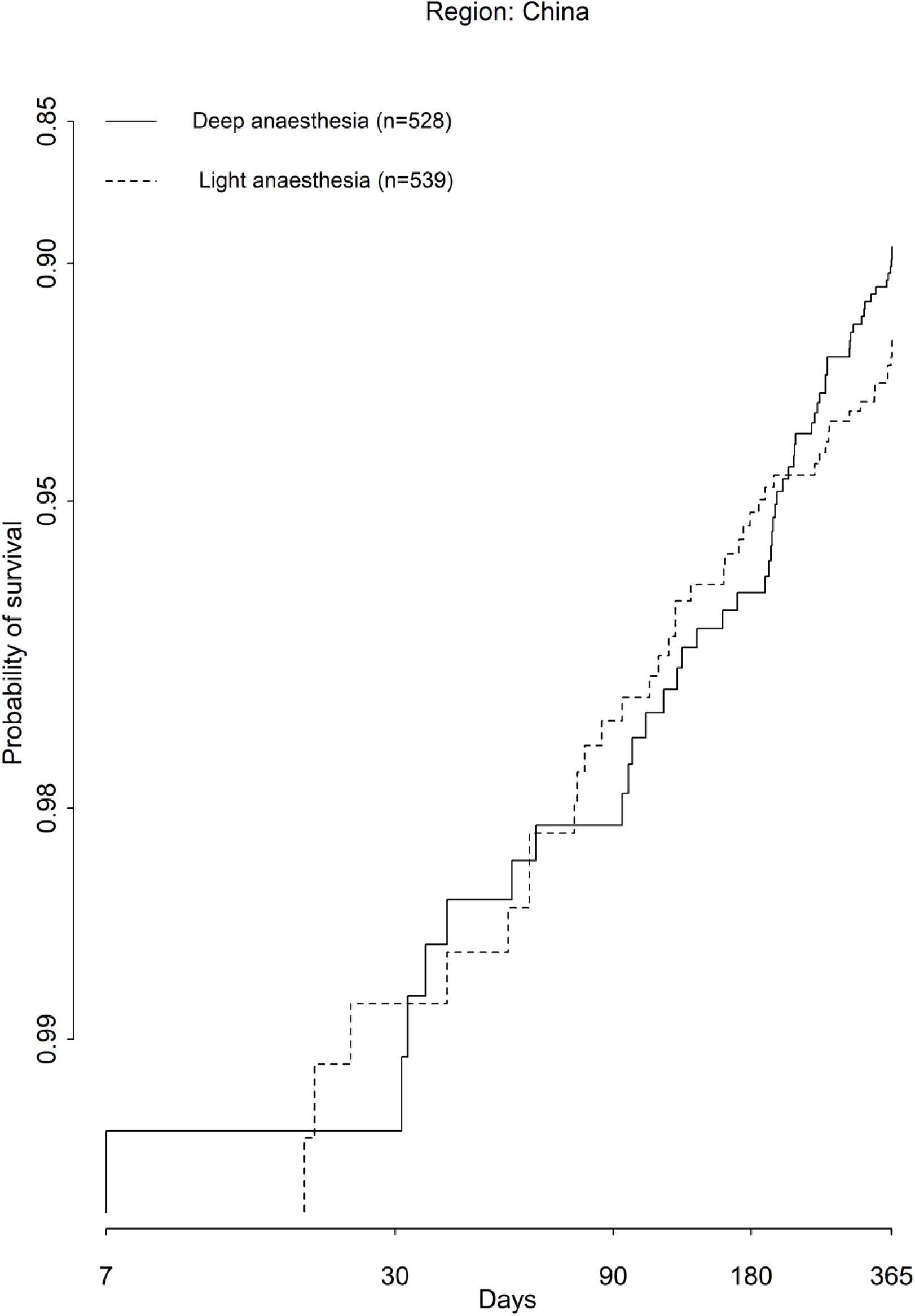
Plot of log (-log(probability of survival)) as a function of log survival time – for patients randomised in China to light (target BIS 50) or deep (target BIS 35) anaesthesia. A proportional hazards ratio would lead to parallel curves for the two randomised groups.

**Figure 3.**
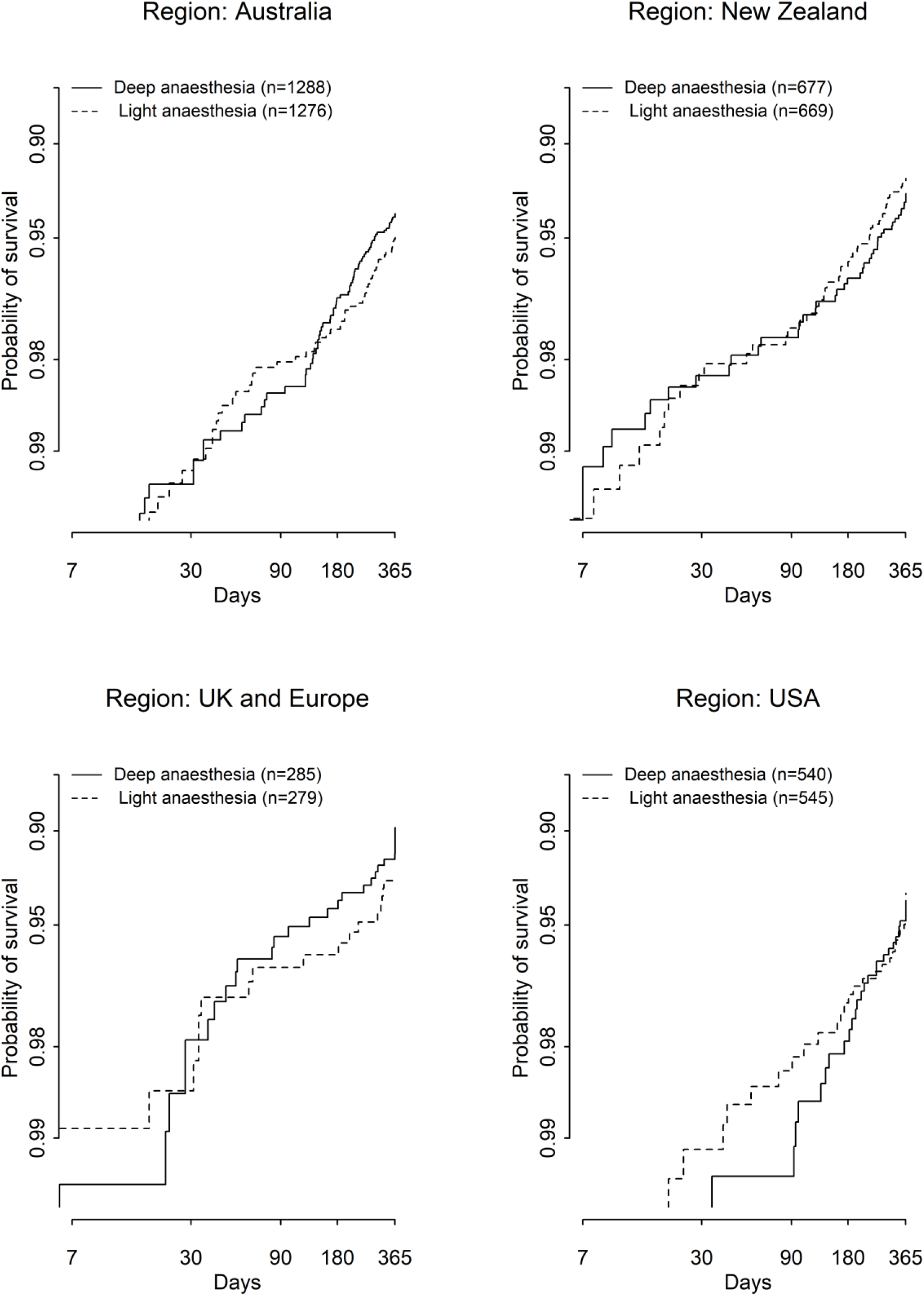
Plots of log (-log(probability of survival)) as a function of log survival time – for patients randomised in regions other than China to light (target BIS 50) or deep (target BIS 35) anaesthesia. A proportional hazards ratio would lead to parallel curves for the two randomised groups.

## Discussion

Our results were unexpected. First, we failed to make more precise estimates of the per-protocol effect of deep anaesthesia. The gain in precision through using all patients for that estimate was offset by the additional variance introduced when modelling missing BIS values. Second, we found that the separation between randomised groups was far closer to the target of 15 BIS units in China than in any other region. Third, we found some evidence that the relative risk of mortality with deep anaesthesia was not constant but increased with time. We had expected the opposite (Appendix A): that mortality unrelated to the intervention would become far more common over time than mortality related to the intervention,^7^ leading to a declining relative risk.

We did however reproduce the original per-protocol estimate using a completely different statistical approach. The original estimate was made by excluding nearly 40% of trial patients, eroding the protection afforded by randomisation and potentially introducing bias if patients considered at greater risk of mortality were further from their target BIS. Our adjustment for potential bias caused no material change to the estimate. We conclude that these estimates are reliable measures of the relative risk of deep versus light anaesthesia on one year mortality. These estimates are consistent with the 20% increase in mortality with deep anaesthesia seen in observational studies (Figure 1). In that sense, there is no controversy.

In the original estimates, increasing the separation from 8.4 to 12.3 BIS units through patient exclusion shifted the hazard ratio from 1.14 to 1.16 (Figure 1). Increasing the separation between randomised groups to 15 BIS units, through covariate adjustment, did not shift the point estimate any further. A possible explanation for this lack of effect is a weak negative correlation between mean achieved BIS and anaesthetic dose. The ‘drug titration paradox’ implies a positive correlation between BIS and anaesthetic dose, instead of the expected negative correlation – increasing the dose is expected to lead to a lower BIS.^24^ In the Balanced trial, the correlation between BIS and dose was at best only weakly negative and in the deep anaesthesia randomised group (Figure 4). Hence a lower BIS was not strongly associated with receipt of a higher dose of anaesthetic.

**Figure 4.**
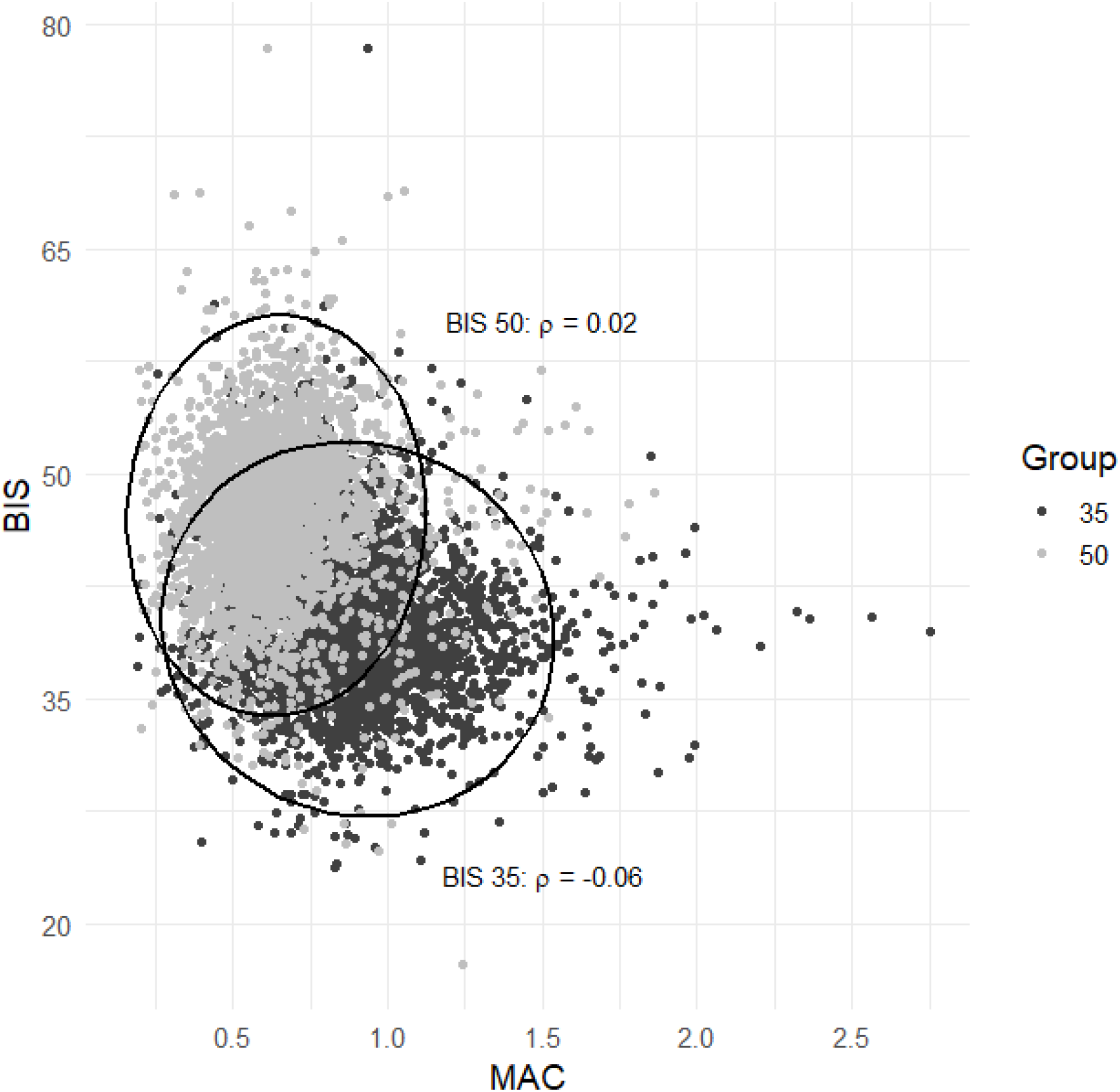
Mean achieved BIS plotted against dose (minimum alveolar concentration, MAC) for patients randomised to either light (target BIS 50) or deep (target BIS 35) anaesthesia. Elipses show a 95% confidence region for each randomised group, assuming a bivariate normal distribution for mean achieved BIS and MAC, and that distribution is summarised by a Pearson correlation coefficient (ρ).

That we expected the hazard ratio to decline with time was perhaps naive. Sub-group analyses of observational data suggest deep anaesthesia might have little or no association with mortality in the first 30 days, but a greater association in studies with follow-up beyond one year.^1^ Our model with two hazard ratios – for events during the first 30 days and one for events after that – gave results consistent with those two sub-group analyses. However, all of these estimates are imprecise – those from observational data and ours from the Balanced trial.

Most of the patients in the delirium sub-study were from China and the BIS separation in that sub-study matches the separation in the Balanced trial for that region (Table 1).^10^ In the sub-study, the incidence of postoperative delirium was higher and one year cognitive function worse under deep anaesthesia,^10^ seemingly driven by a difference in syndromal delirium in Asian patients.^11^ We show a clear crossing of hazards for patients randomised in China, with increasing late mortality under deep anaesthesia (Figure 2). The same pattern is plausible in other regions but the pattern is not clear yet and longer follow up is needed (Figure 3). We hypothesise that greater BIS separation between randomised groups in China led to differences in late mortality between randomised groups in that region that are muted in other regions. The sub-study shows deep anaesthesia led to a greater incidence of postoperative delirium;^10^ delirium has been associated with poor long-term outcomes, cognitive decline and mortality.^25-27^ Results from adjusted meta-analyses of cognitive decline suggest that delirium is not a marker for already compromised cerebral function but that cognitive decline is a consequence of the delirium itself,^25,26^ and that after delirium in surgical patients, the risk of postoperative cognitive impairment increases with time.^26^

It is not clear whether anaesthesia itself can cause delirium in elderly surgical patients.^28^ The aetiology of delirum is multifactorial and several neurobiological pathways are likely to be involved.^29^ Volatile anaesthesia may cause or add to neuroinflammation,^30^ or could be neurotoxic.^31^ But beyond the dose of anaesthesia, there were other differences between randomised groups in the Balanced trial, and in its sub-study, that have been associated with delirium and that could be causal factors. These include the greater use of benzodiazepines and of inotropes or vasopressors and lower mean arterial pressure in those randomised to deep anaesthesia.^32^ Caution about causality is appropriate but new evidence needs to be carefully considered too. The Balanced trial is one of three trial designs identified as potentially informative about whether general anaesthesia leads to cognitive decline.^33^

Clinicians in both randomised groups were seemingly reluctant to sedate patients to target BIS. Clinical equipoise did not require that the individual trial anaesthetists themselves had no preference for one treatment over other alternatives.^34^ While being guided by a ‘precautionary principle’ is understandable, medical research actually requires managed risk.^35^ In clinical practice – as opposed to medical research – when settling on a ‘Goldilocks’ level of sedation, anaesthetists must balance the established risks associated with light anaesthesia with the potential risks associated with deep anaesthesia.^2^ Before the delirium sub-study was published, expert opinion held that it was better to err on the side of deep anaesthesia.^2^ But the delirium sub-study and the results we present here suggest that such an opinion needs re-evaluating, at least when it comes to sedating vulnerable patients.

Like most secondary analyses, our analysis was informed by the primary analysis of these data and by comment on that analysis. Our analysis is therefore exploratory, generating a new hypothesis for further study. Others have made a good case for randomising patients to different doses of anaesthesia in future trials, rather than to different sedation targets.^36,37^ Even so, trial anaesthetists may be uncomfortable with the level of sedation induced by the randomised dose, leading to protocol violations and the need for adjusted per-protocol analyses similar to those in this report. Our results suggest that in future trials, patients should be followed beyond one year and efforts made to ascertain the cause of death. A long-term sub-study was planned for the Balanced trial but did not receive funding. To achieve greater power, larger pragmatic trials in a general population may be necessary, imbedded in routine practice; or alternatively, conventional trials might be targeted at those with the highest risk of delirium.^7^

Ultimately we did not succeed in making more precise estimates of the effect of deep anaesthesia on one year mortality. However, to continue the metaphor of trial by jury,^5^ we found circumstantial evidence that for those randomised to deep anaesthesia in the Balance trial, an increase in the incidence of postoperative delirium led to an increase in late mortality. Deep anaesthesia, in the Balanced trial and in general, consists of a constellation of potentially causal risk factors – a higher dose of general anaesthetic, greater use of benzodiazepines and vasopressors and lower mean arterial pressure. It is not possible to say, with data from the Balanced trial, which of these factors might have increased the incidence of delirium and to what extent.

## Data Availability

Individual, de-identified participant data used in these analyses will be shared by request from any qualified investigator after approval of a protocol, statistical analysis plan, and receipt of a signed data access agreement via the Research Office of
Auckland District Health Board, New Zealand; and after obtaining the approval of the New Zealand Health and Disability Ethics Committees for the project and data release.

## Acknowledgements

The Balanced trial was supported by grants from the Health Research Council of New Zealand (12-308-Short), the Australian National Health and Medical Research Council (APP1042727), the Research Grants Council of Hong Kong (number 61513), the National Institute for Health and Research in the UK (portfolio status), and the National Institutes of Health in the USA (P30 CA 008748). There was no financial support for this secondary analysis.

TS is a consultant to Becton Dickinson (Melbourne, Victoria, Australia) and has received research funding from Boehringer Ingelheim. LAS has received speaker honoraria and research support from Medtronic. JY and SDK declare no competing interests.

## Notes

### Clinical Trial

ACTRN12612000632897

### Funding Statement

This study did not receive any funding. The Balanced trial was supported by grants from the Health Research Council of New Zealand (12-308-Short), the Australian National Health and Medical Research Council (APP1042727), the Research Grants Council of Hong Kong (number 61513), the National Institute for Health and Research in the UK (portfolio status), and the National Institutes of Health in the USA (P30 CA 008748).

### Author Declarations

A protocol for this study and a statistical analysis plan (Appendix A) were approved by the New Zealand Health and Disability Ethics Committees (EXP13559; H Walker, Chair; 15 November 2022) and by the Research Review Committee Te Toka Tumai Auckland (A+9688; M-A Woodnorth, Manager, Research Office; 16 December 2022). Data sharing was subject to a data management plan and a formal signed agreement (12 May 2023).

## References

1 Zorrilla-Vaca A, Healy RJ, Wu CL, Grant MC. Relation between bispectral index measurements of anesthetic depth and postoperative mortality: a meta-analysis of observational studies. Can J Anaesth 2017; 64: 597–607.

2 Fritz BA, Budelier TP, Ben Abdallah A, Avidan MS. The unbearableness of being light. Anesth Analg 2020; 131: 977–980.

3 Short TG, Leslie K, Chan MT, Campbell D, Frampton C, Myles P. Rationale and design of the Balanced Anesthesia Study: a prospective randomized clinical trial of two levels of anesthetic depth on patient outcome after major surgery. Anesth Analg 2015; 121: 357–365.

4 Short TG, Campbell D, Frampton C et al. Anaesthetic depth and complications after major surgery: an international, randomised controlled trial. Lancet 2019; 394: 1907–1914.

5 Galley HF, Webster NR. Deep anaesthesia and poor outcomes: the jury is still out. Lancet 2019; 394: 1881–1882.

6 Charier D, Longrois D, Chapelle C, Salaun JP, Molliex S. Deep anaesthesia and postoperative death: Is the matter resolved? Anaesth Crit Care Pain Med 2020; 39: 21–23.

7 Spence J, Ioannidis JPA, Avidan MS. Achieving balance with power: lessons from the Balanced Anaesthesia Study. Br J Anaesth 2020.

8 Vlisides PE, Ioannidis JPA, Avidan MS. Hypnotic depth and postoperative death: a Bayesian perspective and an independent discussion of a clinical trial. Br J Anaesth 2019; 122: 421–427.

9 Dell-Kuster S, Steiner LA, Young J. Deep anaesthesia. Lancet 2020; 396: 665–666.

10 Evered LA, Chan MTV, Han R et al. Anaesthetic depth and delirium after major surgery: a randomised clinical trial. Br J Anaesth 2021; 127: 704–712.

11 Whitlock EL, Gross ER, King CR, Avidan MS. Anaesthetic depth and delirium: a challenging balancing act. Br J Anaesth 2021; 127: 667–671.

12 Short TG, Leslie K, Campbell D, Frampton C, Chan MTV, Myles PS. Deep anaesthesia - authors’ reply. Lancet 2020; 396: 666–667.

13 Hernan MA, Robins JM. Per-protocol analyses of pragmatic trials. N Engl J Med 2017; 377: 1391–1398.

14 Kahan BC, Jairath V, Dore CJ, Morris TP. The risks and rewards of covariate adjustment in randomized trials: an assessment of 12 outcomes from 8 studies. Trials 2014; 15: 139.

15 Hauck WW, Anderson S, Marcus SM. Should we adjust for covariates in nonlinear regression analyses of randomized trials? Control Clin Trials 1998; 19: 249–256.

16 Bellera CA, MacGrogan G, Debled M, de Lara CT, Brouste V, Mathoulin-Pelissier S. Variables with time-varying effects and the Cox model: some statistical concepts illustrated with a prognostic factor study in breast cancer. BMC Med Res Methodol 2010; 10: 20.

17 Austin PC, Manca A, Zwarenstein M, Juurlink DN, Stanbrook MB. Covariate adjustment in RCTs results in increased power to detect conditional effects compared with the power to detect unadjusted or marginal effects. J Clin Epidemiol 2010; 63: 1391–1393.

18 Schafer JL. 4.5.5 Further comments on imputation modeling. In: Analysis of incomplete multivariate data. Chapman & Hall; 1997 pp. 143–145.

19 Schafer JL. 4.3.2 Inference for a scalar quantity. In: Analysis of incomplete multivariate data. Chapman & Hall; 1997 pp. 107–112.

20 Maheshwari A, McCormick PJ, Sessler DI et al. Prolonged concurrent hypotension and low bispectral index (’double low’) are associated with mortality, serious complications, and prolonged hospitalization after cardiac surgery. Br J Anaesth 2017; 119: 40–49.

21 Xu YX, Shan ZM, Zhao Y, Xiu HH, Xu KQ. Association between depth of anesthesia and postoperative outcome: a systematic review and meta-analysis. Int J Clin Exp Med 2018; 11: 3023–3032.

22 Liu YH, Qiu DJ, Jia L et al. Depth of anesthesia measured by bispectral index and postoperative mortality: a meta-analysis of observational studies. J Clin Anesth 2019; 56: 119–125.

23 Punjasawadwong Y, Chau-In W, Laopaiboon M, Punjasawadwong S, Pin-On P. Processed electroencephalogram and evoked potential techniques for amelioration of postoperative delirium and cognitive dysfunction following non-cardiac and non-neurosurgical procedures in adults. Cochrane Database Syst Rev 2018; 5: CD011283.

24 Schnider TW, Minto CF, Filipovic M. The drug titration paradox: Correlation of more drug with less effect in clinical data. Clin Pharmacol Ther 2021; 110: 401–408.

25 Goldberg TE, Chen C, Wang Y et al. Association of delirium with long-term cognitive decline: A meta-analysis. JAMA Neurol 2020; 77: 1373–1381.

26 Huang H, Li H, Zhang X et al. Association of postoperative delirium with cognitive outcomes: A meta-analysis. J Clin Anesth 2021; 75: 110496.

27 Witlox J, Eurelings LS, de Jonghe JF, Kalisvaart KJ, Eikelenboom P, van Gool WA. Delirium in elderly patients and the risk of postdischarge mortality, institutionalization, and dementia: a meta-analysis. JAMA 2010; 304: 443–451.

28 Aldecoa C, Bettelli G, Bilotta F et al. Update of the European Society of Anaesthesiology and Intensive Care Medicine evidence-based and consensus-based guideline on postoperative delirium in adult patients. Eur J Anaesthesiol 2024; 41: 81–108.

29 Wilson JE, Mart MF, Cunningham C et al. Delirium. Nat Rev Dis Primers 2020; 6: 90.

30 Jin Z, Hu J, Ma D. Postoperative delirium: perioperative assessment, risk reduction, and management. Br J Anaesth 2020; 125: 492–504.

31 Casey CP, Lindroth H, Mohanty R et al. Postoperative delirium is associated with increased plasma neurofilament light. Brain 2020; 143: 47–54.

32 Linassi F, Maran E, Spano L, Zanatta P, Carron M. Anaesthetic depth and delirium after major surgery. Comment on Br J Anaesth 2022; 127: 704–12. Br J Anaesth 2022; 129: e33-e35.

33 Aranake-Chrisinger A, Whitlock EL, Avidan MS. We may be Homo sapiens, but anaesthetists are merely apes when evaluating risk. Br J Anaesth 2018; 121: 702–705.

34 Freedman B. Equipoise and the ethics of clinical research. N Engl J Med 1987; 317: 141–145.

35 Gignon M, Ganry O, Jarde O, Manaouil C. The precautionary principle: is it safe. Eur J Health Law 2013; 20: 261–270.

36 Gaskell A, Sleigh J. The quagmire of postoperative delirium: does dose matter? Br J Anaesth 2021; 127: 664–666.

37 Schnider TW, Minto CF. Beware the drug titration paradox. Comment on Br J Anaesth 2021; 127: 704–12. Br J Anaesth 2022; 128: e335-e337.

