## Appendix A for "Alternative per-protocol estimates: secondary analyses of data from the Balanced randomised controlled trial"

#### Appendix A: Study protocol and analysis plan

Jim Young, Salome Dell-Kuster, Luzius Steiner, Tim Short – third revision 29 September 2022

[In this third revision, material from separate two separate documents – a protocol and an analysis plan – has been combined into this single document and some background material has been removed.]

#### 1. Context

The Balanced trial represents the best data collected to date to answer a question fundamental to general anaesthesia: does greater anaesthetic depth have adverse clinical consequences? The paper describing the trial published in The Lancet contains analyses that are necessary and expected for any randomised clinical trial.[1]

The paper includes a per-protocol analysis presented as a supporting secondary analysis. In that analysis, patients were excluded if their achieved BIS was more than five units from target. This reduced the number of patients from 6626 in the intent-to-treat analysis to 4060, a reduction of nearly 40%. As a result, the estimate of the effect of anaesthetic depth on one year mortality was of a lower precision in this per-protocol analysis than in the intent-to-treat analysis.[1] Excluding these patients increased the separation between treatment groups, from 8.4 to 12.3 BIS units, but the separation was still less than 15 BIS units, the target in this trial.[1]

Per-protocol analyses are preferred in explanatory trials; trials where the focus is on exploring whether an intervention can work (rather than on whether it will work in practice). In a letter to the Lancet, we described a method of analysis that would both allow these excluded patients to be retained in a per-protocol analysis and achieve the target separation of 15 BIS units between the two treatment groups.[2]

As a consequence of our letter, in this study protocol we describe additional per-protocol estimates we think are warranted to better address the fundamental question that motivated the Balanced trial. We provide a detailed analysis plan in which we explain the variables we require – a subset of the data collected in the trial – and what we will do with them. If these additional analyses add to what is known about the clinical consequences of anaesthetic depth, a research letter describing these analyses will be prepared with co-authors from both our study team and the team responsible for the Balanced trial.

#### 2. Background

General anaesthesia has to balance the risk of patients being aware during surgery against the risk of adverse events associated with anaesthesia that is "too deep".[3] Yet it is not clear if there are risks associated with deep anaesthesia: "many anesthesia clinicians now believe dogmatically that deep hypnosis during anesthesia is injurious, and this belief has become entrenched in anesthesiology and critical care lore and literature, despite the lack of robust corroborative evidence and even some compelling contradictory evidence."[3]

The data supporting this belief are largely observational: "the decision to avoid deep anesthesia hypnosis based on these observational data requires one to assume that the observed associations represent causal relationships."[3] The Balanced trial was designed to end the controversy. "This randomized controlled trial should definitively answer the question of whether titrating anesthetic depth makes a difference to patient outcome in a vulnerable patient."[4]

Unfortunately results from this trial have not ended the debate.[5-7] Those commenting have all identified the power of the trial as the key issue. "However, a key methodological consideration that limits interpretation and application of the trial's results is a lack of sufficient statistical power to detect differences in the primary outcome; the sample size was inadequate to answer the trial's primary question."[7]

While this might be seen as a criticism of the Balanced trial, we believe that such criticism is unfair. The Balanced trial was as large as it realistically could have been. Those commenting concede this point when they conclude that approaches other than a conventional randomised clinical trial might be needed to answer the question.[6-8]

But before taking that step, we should first make the best estimates possible with the best data we already have. For this reason, we request data from the Balanced trial in order to make per-protocol estimates that should be more efficient than existing estimates. Essentially we believe we can increase the power of this trial by making more precise estimates. Collecting data is expensive: for little additional effort we may be able to improve on the precision of existing estimates and provide additional insights on the causal question of whether anaesthetic depth effects patient outcome.

#### 3. Explanatory and pragmatic trials

The Balanced trial was designed as an explanatory trial. Key design decisions that illustrate the explanatory intent of the design were the selection of a high risk population (elderly comorbid patients undergoing major surgery), intervention targets and monitoring of those targets with feedback to site coordinators if targets were not routinely met.[9] The aim of the trial was "to definitively answer the question of causality and whether titrating anesthetic depth makes a difference to patient outcome in a vulnerable patient group".[4]

The analysis, however, stressed pragmatism. This can be seen in the choice of a primary outcome important to patients (one-year mortality) and the primary analysis an intent-to-treat analysis that included patients regardless of their achieved BIS.[1] As a consequence, the analysis "tested whether an intervention to control depth within tight, predetermined parameters is effective..."[10]

This is really not the same as answering the causal question of whether anaesthetic depth influences patient outcome.

Per-protocol analyses are more important in explanatory trials.[9] Excluding patients that did not actually adhere to their randomised treatment provides estimates that better measure whether the intervention can work (rather than whether it will work in practice). A per-protocol analysis was carried out and presented as a supporting secondary analysis. Even then, this per-protocol analysis did not reach the target separation between treatment groups of 15 BIS units, and it omitted many patients. It was, therefore, of a lower precision than the intent-to-treat analysis.[1]

The results presented in the Lancet paper are the usual starting point for the analysis of a randomised clinical trial. The results reflect analyses that are necessary and expected. However additional secondary analyses are warranted. In a letter to the Lancet, we described an alternative per-protocol analysis capable of achieving two goals: the precision that comes with including all randomised patients in the analysis and the target 15 BIS unit separation between groups.[2] This alternative analysis has deviation from target BIS as a covariate in any regression model used to estimate the effect of anesthetic depth on primary or secondary outcomes.

At the time we wrote the letter, we did not appreciate the level of missingness in BIS values. This makes our proposed alternative analysis more difficult, although still feasible.

#### 4. Missing BIS values

Not all patients have an electronically recorded mean achieved BIS. A mean achieved BIS value is available for 6445 patients, 97% of the patients in the intent-to-treat population.[1] An analysis of the primary outcome for these 6445 patients suggests that omitting patients with missing BIS values leads to biased estimates, because the hazard ratio for patients with known BIS (0.94, 95% CI 0.77 to 1.14, Short pers comm) is closer to one (no effect) than the hazard ratio for the intent-to-treat population (0.88, 95% CI 0.73 to 1.07 [1]).

The solution is to retain these patients in our analysis, so that we use the same intent-to-treat population as in the original analysis. This mitigates any bias introduced by omitting patients from the intent-to-treat population. However where a patient has a missing BIS value, this missing value needs to be replaced by multiple imputation.[11]

The process of replacing missing BIS values requires a model for imputing BIS from baseline covariates. To properly account for uncertainty in the imputation process, we will be working with multiple data sets for each outcome, each data set with a (randomly selected) imputed plausible value for each missing BIS value. Standard software is then used to estimate a hazard ratio from each of multiple data sets and multiple estimates then combined to produce a final estimate with a confidence interval that reflects both the uncertainty in the original data and the uncertainty in the imputation process.

It is possible that missing BIS values are associated with particular sites within certain countries, rather than with patient characteristics. To allow for this, the site where the patient was recruited will need to be a factor in the imputation model.

#### 5. Non-proportional hazards

When mortality is used as an outcome, it is typically assessed over a long period – a year in the Balanced trial. This may be necessary because the effects of an intervention take time to become apparent. But it can also lead to hazard ratios that change, becoming closer to one as time increases, so the intervention appears to have no effect. This happens if mortality unrelated to the intervention becomes far more common over time than mortality related to the intervention. This is equivalent to a non-proportional hazard; that is, a relative risk that is highest in the period immediately after surgery and which then decays with time.

In a sub-study of the Balanced trial, postoperative delirium was more frequent with deeper anaesthesia – and delirium was associated with subsequent mortality.[12] Non-proportional hazards are a possible explanation for apparently discrepant findings in the parent trial and in the subsequent delirium sub-study. While other explanations are possible,[13] we think it worth assessing evidence for non-proportional hazards in estimates made from these data for the primary outcome.

#### 6. Limitations

There are two main limitations to the additional per-protocol estimates we propose. First, the replacement of missing BIS values assumes that missingness is either random in the first place or can be adequately predicted by an imputation model so that the process of imputation leads to unbiased estimates of missing BIS values. Second, mean achieved BIS is measured after randomisation and there is the possibility of confounding between BIS deviation from target and outcome (as in an observational study). The solution to this problem is to include baseline covariates from the imputation model in the outcome model where those covariates could be associated with both BIS deviation from target and mortality (as in an observational study).

Essentially what we are proposing is an observational analysis of data collected in a randomised trial. This is because we assume that both (1) we can make unbiased estimates of missing BIS values from baseline covariates and (2) adding additional baseline covariates will be sufficient to adjust for any confounding that arises from including a covariate in outcome models that was measured after randomisation (deviation from target BIS).

That said, the original per-protocol estimate is also at risk of an equivalent bias. Nearly 40% of patients in the intent-to-treat population were excluded from the original per-protocol analysis, again reducing the protection provided by randomisation. Those patients excluded – because their achieved BIS was more than five units from target – are potentially different from those with achieved BIS close to target. These excluded patients might have been assessed by the anaesthetist as more at risk of an adverse event and then their exclusion creates a bias.

### 7. Analysis plan

#### 7.1. Summary

In this analysis plan, we describe the additional analyses that we think have value. These analyses should provide more efficient per-protocol estimates of the effect of the treatment group on the primary outcome and on secondary outcomes. For each additional analysis, we describe and explain the statistical methods we will use and list the variables we will need. All analyses will be carried out in SAS 9.4.

The variables needed are summarised in Table 1. These variables are: baseline characteristics (including age at surgery in years and sex), treatment group and mean achieved BIS, region and site within region; the primary outcome (survival up to one year postoperative) and seven secondary outcomes. For each variable, we need all available values for the 6626 patients in the intent-to-treat population.

No additional data are needed beyond those already collected in the Balanced trial.

The analysis plan below contains technical information, suitable for review by an experienced statistician. The analyses proposed can be summarised as:

1. Reproducing the original estimates of the effect of the treatment group on the primary outcome (one year mortality) in the original intent-to-treat and per-protocol trial populations. These estimates are hazard ratios from Cox regression models.

2. A new per-protocol estimate for the primary outcome (one year mortality) using the original intent-to-treat population but with an additional covariate – deviation from target BIS – in the Cox regression model. To make this estimate, we will need to impute mean achieved BIS, where it is missing, by multiple imputation. Mean achieved BIS is missing for 3% of the intent-to-treat population. The estimated hazard ratio from this model should be more precise than the original per-protocol estimate and using this model achieves the target separation of 15 BIS units between the two treatment groups.

3. A new per-protocol estimate for the primary outcome (one year mortality) using the original intent-to-treat population but with two additional covariates in the Cox regression model: deviation from target BIS (as above) and either age at surgery or Charlson comorbidity index at surgery (because this takes into account both age and comorbidity). Adding the second additional covariate may further improve precision.

4. A new per-protocol estimate for the primary outcome (one year mortality) using the original intent-to-treat population but with three or four additional covariates in the Cox regression model: deviation from target BIS (as above) and the two or three covariates found to be most predictive of mean achieved BIS in imputation modelling. Mean achieved BIS can be thought of as a measure of adherence to the trial protocol. There is the possibility of confounding between BIS deviation from target and mortality (as in an observational study). The solution to this

problem is to include baseline covariates in the Cox regression model that were used in the imputation model if strongly associated with both mean achieved BIS and mortality.

5. A new per-protocol estimate for the primary outcome (one year mortality) using the original intent-to-treat population but with two additional covariates in the Cox regression model: deviation from target BIS (as above) and a time dependent term representing an interaction between treatment group and log time. The estimated hazard ratio for this second covariate can be used to assess whether the effect of anaesthetic depth on mortality changes with time (a non-proportional hazard).

6. New per-protocol estimates for seven secondary outcomes (listed in Table 1) using the original intent-to-treat population but with an additional covariate – deviation from target BIS (as above) – in the logistic regression model for each outcome.

##### 7.2. Reproducing original estimates

The first step in any further analysis is to reproduce original estimates to make sure we understand the data we receive.

The original intent-to-treat estimate was made "from a Cox regression model, which included randomised treatment and region as factors. Patients who were lost to follow-up were censored to the last time that they were known to be alive after hospital discharge."[1]

To reproduce this estimate therefore requires four variables: randomised treatment, region, follow up time, and an event indicator (0=censored, 1=died); each variable with a value for each patient in the intent-to-treat analysis (n=6626). To reproduce the original per-protocol estimate requires an additional indicator variable with value one for those patients included in the per-protocol population.

##### 7.3. Deviation from BIS target as a covariate

In our letter to the Lancet, we describe the advantages of adding an extra covariate to intent-to-treat analyses where that covariate represents each patient's deviation from their BIS target (35 or 50 depending on their randomised treatment).[2] This gives a per-protocol estimate using all patients – so there is no loss of power – and, if a lower BIS is associated with increased mortality, adding this covariate will even increase the power of the study.[14] That is, these alternative per-protocol estimates will be more precise, with narrower confidence intervals, than the original per-protocol estimates.

There is also a risk of bias in the original per-protocol estimates because nearly 40% of patients in the intent-to-treat population were excluded from per-protocol analyses. Those patients excluded – because their achieved BIS was more than five units from target – are potentially different from those with achieved BIS close to target. These patients might have been assessed by the anaesthetist as more at risk of an adverse event and then their exclusion creates a selection bias.[15] Our alternative estimates retain all patients in the intent-to-treat population and this avoids any selection bias.

However, more importantly, estimates from this alternative approach will be conditional on a 15 unit BIS separation between treatment groups – reproducing the target difference between treatments the trial was designed to evaluate. In the trial, "targets were chosen on the basis of previous published research, audit data from a large hospital database, where these targets were close to the first and third quartiles of mean BIS in a similar group of patients, and the manufacturer's recommendations for appropriate targets for general anaesthesia."[1]

To carry out these alternative analyses requires a single additional variable for each patient in the intent-to-treat analysis: the patient's mean achieved BIS.

##### 7.4. Multiple imputation of missing mean achieved BIS

Mean achieved BIS is missing for 3% of the patients in the intent-to-treat population.[1] We will replace missing BIS values using multiple imputation so that patients with a missing BIS value can be retained in our analyses. Note that multiple imputation implies that we will be working with multiple datasets for each outcome, each dataset with each missing BIS replaced by a (randomly selected) imputed plausible value. Standard software is then used to estimate a hazard ratio from each of these multiple datasets and multiple estimates are then combined to produce a final estimate with a confidence interval that reflects both the uncertainty in the original data and the uncertainty in the imputation process.[11]

Our imputation model will be based on sequential regressions where variables are arranged in sequence to achieve a monotone pattern of missingness.[16] In brief, we will estimate values for a nearly complete variable such as the preoperative Charlson comorbidity index from values of fully complete variables such as treatment group, age, sex and indicators for coexisting stroke or neurological disease and coexisting cardiovascular disease. We will then estimate values of deviation from target BIS from nearly complete and fully complete variables. We may try a variety of multiple imputation models, once we see the pattern of missingness in these data, adding additional variables to the model that are either associated with deviation from target BIS (such as the preoperative WHODAS 2.0 score) or with the pattern of missingness (such as indicators denoting regions or sites within regions with frequent missing BIS values).[17] It is possible that missing BIS values are associated with particular sites within certain countries, rather than with patient characteristics. To allow for this, the site where the patient was recruited will need to be a factor in the imputation model.

We will judge the suitability of candidate imputation models using variance statistics; in particular the between imputation variance for deviation from target BIS. We will not carry out any analyses of outcomes until we have selected a preferred imputation model.

Our analyses of outcomes will use 200 datasets with, in each dataset, each missing BIS replaced by a randomly selected imputed value.[11] With modern computing power, there is no real disadvantage in using more datasets than might be strictly necessary.

For our imputation modelling, we need the following variables: the patient's age at surgery (in years), their preoperative Charlson comorbidity index, their preoperative WHODAS 2.0 score, indicators for coexisting stroke or neurological disease and for coexisting cardiovascular disease, and a code representing the site within the region where the operation took place.

##### 7.5. Age as a covariate and as a potential effect modifier

Analyses of randomised trials typically do not use prognostic covariates. However including appropriate covariates will increase the precision of estimates of the treatment effect, even in non-linear models such as the Cox model.[18] It is likely that one year mortality is strongly associated with age at surgery; therefore including age at surgery as an additional covariate can be expected to lead to estimates with narrower confidence intervals.

Note that in a non-linear model, adding covariates changes the treatment effect being estimated. Adding covariates moves estimates closer to those from a subject-specific model; a model that has a random effect for each patient. "There is no unique population-averaged treatment effect. Every choice of a set of covariates, including the choice of no covariates, is a different population-averaged model."[18]

In an analysis without covariates, estimates are appropriate for patient randomly selected from the trial regardless of their covariates values.[14] In an analysis with BIS deviation from target as a covariate, estimates are appropriate for patients with a 15 unit separation in BIS regardless of other covariate values. In an analysis with both age and BIS deviation from target as covariates, estimates are appropriate for patients of a certain age and a 15 unit separation in BIS regardless of other covariate values. We would centre age around the mean for the trial – age 72 – so that when age is added as a covariate, estimates are appropriate for patients with the average age.

In an exploratory analysis, we would repeat this process replacing patient's age at surgery with the value of the patient's Charlson comorbidity index at surgery. This index takes both the patient's age and co-morbidity into account and was designed to predict one year mortality.[19,20] It might therefore prove to be a better covariate than age alone. The precision in the per-protocol estimate of the treatment effect will be used to assess which is the better covariate – age or the Charlson comorbidity index. With the best covariate, we will then use simulation to make an estimate of the treatment effect conditional only on a 15 unit BIS separation between treatment groups.[21]

Finally, if there is evidence of a treatment effect, it would be appropriate to see whether the effect might vary with age. This requires adding an interaction term to the Cox model – the interaction between treatment group and age. We stress: this would be an exploratory analysis because the trial was not powered to estimate the effect of treatment modifiers. However an additional analysis of this sort might be valuable for future trial design.[22]

To carry out these alternative analyses requires two additional variables for each patient in the intent-to-treat analysis: the patient's age at surgery (in years) and the value of their Charlson comorbidity index prior to surgery. These two variables are also needed for imputation modelling.

##### 7.6. Mean achieved BIS as a measure of adherence

With multiply imputed data, the model used to estimate the effect of an outcome often contains the same variables used to impute missing values. However there are exceptions to this general rule and in the setting of a randomised trial, the outcome model may omit variables used in the imputation model in order to make estimates that are not conditional on these variables.[11] Nevertheless, it would be sensible to add some of the variables in the imputation model to an outcome model, where these variables are potentially associated with both the BIS deviation from target and the outcome.

This is because mean achieved BIS can be thought of as a measure of adherence to the trial protocol. From this perspective, including the BIS deviation from target in a regression model for an outcome could erode the protection from confounding afforded by randomisation. Because mean achieved BIS is measured after randomisation, there is the possibility of confounding between BIS deviation from target and outcome (as in an observational study). The solution to this problem is to include baseline covariates in the outcome model that were used in the imputation model if they could be associated with both BIS deviation from target and outcome – see Panel B of Figure 2 in Hernan and Robins 2017.[15] Again, with additional covariates in the model, we can use simulation to make an estimate of the treatment effect conditional only on a 15 unit BIS separation between treatment groups.[21]

No additional variables are needed for this, beyond those needed for imputation modelling.

##### 7.7. Non-proportional hazards

In a subset of trial patients, postoperative delirium was more frequent with deeper anaesthesia and delirium was associated with subsequent mortality. Non-proportional hazards are a possible explanation for apparently discrepant findings in the parent trial and in the subsequent delirium sub-study.

We will assess non-proportional hazards for the primary outcome in two ways.[23,24] First we will make a visual assessment, plotting log negative log survival against log time and Schoenfeld residuals against time. Second, we will add a time dependent term to the Cox model representing an interaction between treatment group and log time. The parameter estimate for this term provides evidence for whether the hazard ratio increases or decreases with time. Only a decrease with time is plausible.

No additional variables are needed for this.

##### 7.8. Secondary outcomes

In our letter, we note that it is important to look for consistency between estimates for the primary outcome and estimates for important secondary outcomes.[2] We identified the following as important secondary outcomes: myocardial infarction, cardiac arrest, pulmonary embolism, stroke, sepsis, surgical site infection, and unplanned ICU admission.

For each secondary outcome, we would make an estimate adding a single covariate - patient's mean achieved BIS minus their target BIS – to a conditional logistic regression model stratified by region with randomised treatment as the other independent variable. A conditional logistic regression model gives an estimate that is the multivariate equivalent to the Mantel–Haenszel estimate of the common odds ratio originally used for intent-to-treat analyses of secondary outcomes.[25]

These additional analyses require seven secondary outcomes (see Table 1): for each outcome, we need a binary indicator with values for each patient in the intent-to-treat analysis.

#### 8. Own research

Jim Young has been employed as a biostatistician in university research groups in Switzerland and Canada for the last twenty years. He has been entrusted to store and carry out analyses of multinational data collected in both clinical trials [26] and observational cohorts.[27,28] Prior to that, he worked for New Zealand's official statistics agency, Statistics New Zealand, and subsequently on research funded by the agency into data confidentiality.[29,30]

Salome Dell-Kuster is a board-certified anaesthesiologist, and a senior consultant at the University Hospital Basel, Switzerland. She is also a research scientist with formal training in statistics and research methodology.[31] Her research over the last decade has focused on perioperative patient safety and outcome.[32,33] Most recently, she was the chief investigator of an international multicentre cohort study including over 2500 patients in 18 centres from 12 countries, assessing the validity of a novel classification of intraoperative adverse events.[34]

Professor Luzius A. Steiner is the Chairman of the Clinic for Anaesthesia, Intermediate Care, Prehospital Emergency Medicine and Pain Therapy at the University Hospital of Basel, Switzerland. In addition, and since 2021, he has been promoted to the role of Medical Head of the Department of Acute Medicine. His research focuses on postoperative delirium and cognitive dysfunction and the role of perioperative management, cerebral perfusion, and neuromonitoring in the development and prevention of these complications.[35-38] He has lasting strong collaborations with the Neurocritical Care Unit, the Wolfson Brain Imaging Centre, and Academic Neurosurgery at Addenbrooke's Hospital in Cambridge, UK, an affiliate of the University of Cambridge, where he received his PhD for research on cerebral perfusion after head injury.

#### Table 1: Estimates and the data need to make them

| Estimate | Variables needed – with values for all patients in the intent-to-treat analysis (n = 6626) |
|---|---|
| Primary outcome | (1 year mortality) |
| Original intent-to-treat | Randomised treatment<br>Region<br>Site within region*<br>Follow up time<br>Event indicator (0=censored, 1=died) |
| Original per-protocol | Per-protocol indicator (0=excluded, 1=included) |
| Alternative per-protocol | Patient's mean achieved BIS<br>Sex<br>Age at surgery (years)<br>Preoperative Charlson comorbidity index<br>Preoperative WHODAS 2.0 score<br>Coexisting stroke or neurological disease indicator<br>Coexisting cardiovascular disease indicator |
| Secondary outcomes | (All binary indicators at 1 year: 0=no, 1=yes) |
| | Myocardial infarction<br>Cardiac arrest<br>Pulmonary embolism<br>Stroke<br>Sepsis<br>Surgical site infection<br>Unplanned ICU admission |

* Site within region: To improve imputation model stability, it will be useful to combine those sites recruiting only a few patients or group sites according to a sensible classification (such as university, specialist, regional or local hospital). To be discussed.

#### 9. References

1 Short TG, Campbell D, Frampton C, et al. Anaesthetic depth and complications after major surgery: an international, randomised controlled trial. *Lancet* 2019; **394**: 1907-14.

2 Dell-Kuster S, Steiner LA, Young J. Deep anaesthesia. *Lancet* 2020; **396**: 665-66.

3 Fritz BA, Budelier TP, Ben Abdallah A, Avidan MS. The unbearableness of being light. *Anesth Analg* 2020; **131**: 977-80.

4 Short TG, Leslie K, Chan MT, Campbell D, Frampton C, Myles P. Rationale and design of the Balanced Anesthesia Study: a prospective randomized clinical trial of two levels of anesthetic depth on patient outcome after major surgery. *Anesth Analg* 2015; **121**: 357-65.

5 Galley HF, Webster NR. Deep anaesthesia and poor outcomes: the jury is still out. *Lancet* 2019; **394**: 1881-82.

6 Charier D, Longrois D, Chapelle C, Salaun JP, Molliex S. Deep anaesthesia and postoperative death: Is the matter resolved? *Anaesth Crit Care Pain Med* 2020; **39**: 21-23.

7 Spence J, Ioannidis JPA, Avidan MS. Achieving balance with power: lessons from the Balanced Anaesthesia Study. *Br J Anaesth* 2020.

8 Vlisides PE, Ioannidis JPA, Avidan MS. Hypnotic depth and postoperative death: a Bayesian perspective and an independent discussion of a clinical trial. *Br J Anaesth* 2019; **122**: 421-27.

9 Loudon K, Treweek S, Sullivan F, Donnan P, Thorpe KE, Zwarenstein M. The PRECIS-2 tool: designing trials that are fit for purpose. *BMJ* 2015; **350**: h2147.

10 Short TG, Leslie K, Campbell D, Frampton C, Chan MTV, Myles PS. Deep anaesthesia - authors' reply. *Lancet* 2020; **396**: 666-67.

11 Kenward MG, Carpenter J. Multiple imputation: current perspectives. *Stat Methods Med Res* 2007; **16**: 199-218.

12 Evered LA, Chan MTV, Han R, et al. Anaesthetic depth and delirium after major surgery: a randomised clinical trial. *Br J Anaesth* 2021; **127**: 704-12.

13 Whitlock EL, Gross ER, King CR, Avidan MS. Anaesthetic depth and delirium: a challenging balancing act. *Br J Anaesth* 2021; **127**: 667-71.

14 Kahan BC, Jairath V, Dore CJ, Morris TP. The risks and rewards of covariate adjustment in randomized trials: an assessment of 12 outcomes from 8 studies. *Trials* 2014; **15**: 139.

15 Hernan MA, Robins JM. Per-protocol analyses of pragmatic trials. *N Engl J Med* 2017; **377**: 1391-98.

16 Yuan Y. Multiple Imputation Using SAS Software. *J Stat Soft*. https://www.jstatsoft.org/article/view/v045i06

17 van Buuren S, Boshuizen HC, Knook DL. Multiple imputation of missing blood pressure covariates in survival analysis. *Stat Med* 1999; **18**: 681-94.

18 Hauck WW, Anderson S, Marcus SM. Should we adjust for covariates in nonlinear regression analyses of randomized trials? *Control Clin Trials* 1998; **19**: 249-56.

19 Charlson ME, Pompei P, Ales KL, MacKenzie CR. A new method of classifying prognostic comorbidity in longitudinal studies: development and validation. *J Chronic Dis* 1987; **40**: 373-83.

20 Quan H, Li B, Couris CM, et al. Updating and validating the Charlson comorbidity index and score for risk adjustment in hospital discharge abstracts using data from 6 countries. *Am J Epidemiol* 2011; **173**: 676-82.

21 US Food and Drug Administration. Adjusting for covariates in randomized clinical trials for drugs and biological products: Draft guidance for industry. *FDA-2019-D-0934*. https://www.fda.gov/regulatory-information/search-fda-guidance-documents/adjusting-covariates-randomized-clinical-trials-drugs-and-biological-products

22 Kraemer HC, Frank E, Kupfer DJ. Moderators of treatment outcomes: clinical, research, and policy importance. *JAMA* 2006; **296**: 1286-89.

23 Schemper M. Cox analysis of survival data with non-proportional hazard functions. *The Statistician* 1992; **41**: 455-65.

24 Bellera CA, MacGrogan G, Debled M, de Lara CT, Brouste V, Mathoulin-Pelissier S. Variables with time-varying effects and the Cox model: some statistical concepts illustrated with a prognostic factor study in breast cancer. *BMC Med Res Methodol* 2010; **10**: 20.

25 Agresti A, Min Y. Effects and non-effects of paired identical observations in comparing proportions with binary matched-pairs data. *Stat Med* 2004; **23**: 65-75.

26 Young J, De Sutter A, Merenstein D, et al. Antibiotics for adults with clinically diagnosed acute rhinosinusitis: a meta-analysis of individual patient data. *Lancet* 2008; **371**: 908-14.

27 Young J, Psichogiou M, Meyer L, et al. CD4 cell count and the risk of AIDS or death in HIV-Infected adults on combination antiretroviral therapy with a suppressed viral load: a longitudinal cohort study from COHERE. *PLoS Med* 2012; **9**: e1001194.

28 Klein MB, Young J, Althoff KN, et al. Are modern antiretrovirals hepatotoxic? Signals in patients starting ART in NA-ACCORD. *28th Conference on Retroviruses and Opportunistic Infections, 6-10 March 2021*. https://www.croiconference.org/abstract/are-modern-antiretrovirals-hepatotoxic-signals-in-patients-starting-art-in-na-accord/ (accessed Jun 14, 2021).

29 Young J, Graham P, Penny R. Using Bayesian networks to create synthetic data. *Journal of Official Statistics* 2009; **25**: 549-67.

30 Graham P, Young J, Penny R. Multiply imputed synthetic data: evaluation of hierarchical Bayesian imputation models. *Journal of Official Statistics* 2009; **25**: 245-68.

31 Dell-Kuster S, Droeser RA, Schafer J, et al. Systematic review and simulation study of ignoring clustered data in surgical trials. *Br J Surg* 2018; **105**: 182-91.

32 Dell-Kuster S, Hoesli I, Lapaire O, et al. Efficacy and safety of carbetocin given as an intravenous bolus compared with short infusion for Caesarean section - double-blind, double-dummy, randomized controlled non-inferiority trial. *Br J Anaesth* 2017; **118**: 772-80.

33 Rosenthal R, Hoffmann H, Clavien PA, Bucher HC, Dell-Kuster S. Definition and Classification of Intraoperative Complications (CLASSIC): Delphi Study and Pilot Evaluation. *World J Surg* 2015; **39**: 1663-71.

34 Dell-Kuster S, Gomes NV, Gawria L, et al. Prospective validation of classification of intraoperative adverse events (ClassIntra): international, multicentre cohort study. *BMJ* 2020; **370**: m2917.

35 Hollinger A, Siegemund M, Goettel N, Steiner LA. Postoperative delirium in cardiac surgery: An unavoidable menace? *J Cardiothorac Vasc Anesth* 2015; **29**: 1677-87.

36 Siegemund M, Steiner LA. Postoperative care of the neurosurgical patient. *Curr Opin Anaesthesiol* 2015; **28**: 487-93.

37 Goettel N, Burkhart CS, Rossi A, et al. Associations between impaired cerebral blood flow autoregulation, cerebral oxygenation, and biomarkers of brain injury and postoperative cognitive dysfunction in elderly patients after major noncardiac surgery. *Anesth Analg* 2017; **124**: 934-42.

38 Steiner LA. Postoperative delirium guidelines: The greater the obstacle, the more glory in overcoming it. *Eur J Anaesthesiol* 2017; **34**: 189-91.
