## Appendix B for "Alternative per-protocol estimates: secondary analyses of data from the Balanced randomised controlled trial"

### Appendix B: Analyses of secondary outcomes

Jim Young [1] *, Tim Short [2,3], Luzius A Steiner [4,5], Salome Dell-Kuster [4,5]

[1] Department of Epidemiology, Biostatistics and Occupational Health, Faculty of Medicine, McGill University, Montreal, Canada.

[2] Department of Anaesthesia and Perioperative Medicine, Auckland City Hospital, Auckland, New Zealand.

[3] Department of Anaesthesia, University of Auckland, Auckland, New Zealand.

[4] Clinic for Anaesthesiology, Intermediate Care, Prehospital Emergency Medicine and Pain Therapy, Department of Acute Medicine, University Hospital of Basel, Basel, Switzerland.

[5] Department of Clinical Research, University of Basel, Basel, Switzerland.

* Corresponding Author: Jim Young, Research Institute of the McGill University Health Centre, 5252 boul de Maisonneuve W, #3C.23, Montréal, QC H4A 3S5 Canada. Tel. +1-514-934-1934 ext.32198,

### 1. Introduction

In a letter to The Lancet, we suggested an alternative per-protocol estimate for the effect of deep anaesthesia on one year mortality that might improve the power of the Balanced trial.[1] We also noted the importance of looking for consistency between the estimate for the primary outcome and estimates for important secondary outcomes. We identified the following as important secondary outcomes: myocardial infarction, cardiac arrest, pulmonary embolism, stroke, sepsis, surgical site infection, and unplanned ICU admission.

In this appendix, we report alternative estimates for these seven secondary outcomes. Each estimate has been adjusted by deviation from target BIS, recreating in a statistical sense the desired separation between randomised groups of 15 BIS units.

### 2. Methods

We followed the same process as described for the primary outcome. Again our population of interest was the intent-to-treat population of the Balanced trial. We used this population to make a per-protocol estimate for each secondary outcome by adding an additional covariate – each patient's deviation from target BIS (35 or 50 depending on their randomised group) – to the original intent-to-treat analysis.

For each secondary outcome, we added this covariate to a conditional logistic regression model stratified by region with randomised treatment as the other independent variable. A conditional logistic regression model gives an estimate that is the multivariate equivalent to the Mantel–Haenszel estimate of the common odds ratio originally used for intent-to-treat analyses of secondary outcomes.[2] We used the same sample of 200 multiply imputed data sets as before and Rubin's rule for combining point estimates from multiple imputed data sets.[3]

These secondary analyses were carried out in SAS 9.4 TS Level 1M5 (procedures LOGISTIC and MIANALYZE). We report odds ratios (OR) with light anaesthesia as the reference, as we do with the primary outcome and in contrast to the original analysis.

### 3. Results

A variety of estimates were made for each of these outcomes (Table 1). Estimates in the first two columns reproduce the original estimates with the data we received. These estimates, when inverted, match those in Table 3 of the original Lancet paper and Table 4 of its supplementary appendix.[4] Estimates in the third column were made by conditional logistic regression with the regions as strata. This shows the equivalence between conditional logistic regression and the original method of analysis – the Mantel-Haenszel estimate of a common odds ratio. We needed to use conditional logistic regression in order to add an additional covariate to the analysis – deviation from target BIS. We then combined adjusted point estimates made from multiply imputed data sets and this process provided the final set of adjusted estimates in the fourth column.

**Table 1**: Intent-to-treat (ITT) and per-protocol (PP) estimates of the effect of deep anaesthesia on seven secondary outcomes.

| | Odds ratio (95% confidence interval) with light anaesthesia as the reference | | | |
|---|---|---|---|---|
| Secondary outcome | Mantel-Haenszel stratified by region | | Conditional logistic regression stratified by region | |
| | ITT n=6626 | PP n=4060 | PP unadjusted n=4060 | PP adjusted n=6626 |
| Myocardial infarction | 1.00 (0.73 to 1.38) | 1.28 (0.86 to 1.90) | 1.28 (0.86 to 1.90) | 1.07 (0.89 to 1.30) |
| Cardiac arrest | 0.52 (0.26 to 1.04) | 0.53 (0.22 to 1.27) | 0.54 (0.23 to 1.27) | 0.80 (0.52 to 1.23) |
| Pulmonary embolism | 1.30 (0.83 to 2.05) | 1.32 (0.72 to 2.42) | 1.32 (0.72 to 2.42) | 1.06 (0.79 to 1.42) |
| Stroke | 0.76 (0.48 to 1.20) | 0.98 (0.56 to 1.72) | 0.98 (0.56 to 1.72) | 0.97 (0.73 to 1.29) |
| Sepsis | 1.08 (0.88 to 1.31) | 1.35 (1.04 to 1.74) | 1.35 (1.04 to 1.73) | 1.06 (0.94 to 1.20) |
| Surgical site infection | 0.87 (0.72 to 1.05) | 0.94 (0.73 to 1.21) | 0.94 (0.73 to 1.21) | 0.95 (0.85 to 1.08) |
| Unplanned ICU admission | 1.12 (0.90 to 1.39) | 1.21 (0.92 to 1.60) | 1.21 (0.92 to 1.60) | 1.08 (0.95 to 1.23) |

### 4. Discussion

Of the seven outcomes in Table 1, the first four occurred with low frequency, especially in the per-protocol population (n=4060). This is likely to lead to instability when comparing estimates made from populations of differences sizes (n=6626 or n=4060). With pulmonary embolism, the conditional logistic regression algorithm did not convergence for the per-protocol population. Even without convergence issues, estimates are not going to be very reliable when there are few events. This applies both to the estimate of the effect of deep anaesthesia and to the estimate of the association with deviation from target BIS.

With the primary outcome, the estimated hazard ratio for deviation from target BIS was 0.95 (95% confidence interval, CI, 0.79 to 1.14). This implied that an increasing positive deviation (mean achieved BIS minus target BIS) reduced risk. Because patients in the light anaesthesia group were, on average, below target BIS and patients in the deep anaesthesia group were above target BIS, adjustment tended to reduce the risk for patients under light anaesthesia and increase the risk for patients under deep anaesthesia; the ratio of the two risks then increased (albeit only slightly). With rare events like cardiac arrest and stroke, the estimated odds ratio for deviation from target BIS was lower (respectively OR 0.75, 95% CI 0.39 to 1.45; OR 0.75, 95% CI 0.48 to 1.19) and imprecise compared to its estimate for the primary outcome. This led to an exaggerated increase in the adjusted odds ratio compared to its ITT estimate. So for these first four outcomes, adjustment for deviation from target BIS could not improve on an already unreliable estimate. Estimates of the association with target BIS were imprecise, and lower than the estimate for the primary outcome, except the estimate for pulmonary embolism (which was above one – OR 1.19, 95% CI 0.77 to 1.83).

The remaining three outcomes were relatively frequent – sepsis, surgical site infection, unplanned ICU admission. All three outcomes led to estimates of the association with deviation from target BIS that were almost identical to the estimate for the primary outcome. All three were relatively short-term outcomes, mostly occurring in the first 30 days if they were going to occur at all. The lack of any effect of deep anaesthesia in these outcomes was consistent with the apparent lack of any effect of deep anaesthesia on mortality within the first 30 days.

### 5. References

1 Dell-Kuster S, Steiner LA, Young J. Deep anaesthesia. *Lancet* 2020; **396**: 665-66.

2 Agresti A, Min Y. Effects and non-effects of paired identical observations in comparing proportions with binary matched-pairs data. *Stat Med* 2004; **23**: 65-75.

3 Schafer JL. 4.3.2 Inference for a scalar quantity. *Analysis of incomplete multivariate data*. Chapman & Hall; 1997 pp 107-12.

4 Short TG, Campbell D, Frampton C, et al. Anaesthetic depth and complications after major surgery: an international, randomised controlled trial. *Lancet* 2019; **394**: 1907-14.
